# Association of Heart Failure Subtypes and Atrial fibrillation: Data from the Atherosclerosis Risk in Communities (ARIC) Study

**DOI:** 10.1101/2021.03.30.21254622

**Authors:** Miriam A.M. Nji, Scott D. Solomon, Lin Yee Chen, Amil M. Shah, Elsayed Z. Soliman, Aniqa Alam, Vinita Subramanya, Alvaro Alonso

## Abstract

**Aims:** To determine the prevalence and incidence of AF among HF subtypes in a biracial community-based cohort.

**Methods:** We studied 6,496 participants in the Atherosclerosis Risk in Community study (mean age, 75.8±5.3, 59% women, 23% black) who attended the 2011-2013 visit. HF was identified from physician adjudicated diagnosis, hospital discharges, and self-report. HF subtypes were based on echocardiography. A left ventricular ejection fraction <40% represents HF with reduced ejection fraction (HFrEF), 40%-49% for HF with midrange ejection fraction (HFmEF), and ≥50% for HF with preserved ejection fraction (HFpEF). AF was ascertained through 2017 from study electrocardiograms, hospital discharges, and death certificates. Confounder-adjusted logistic regression and Cox models were used to estimate associations of HF subtype with prevalent and incident AF.

**Results:** Among eligible participants, 393 had HF (HFpEF=232, HFmEF=41, HFrEF=35 and unclassified HF =85) and 735 had AF. Compared to those without HF, all HF subtypes were more likely to have prevalent AF [odds ratio (95% confidence interval (CI)) 7.4 (5.6-9.9) for HFpEF, 8.1 (4.3-15.3) for HFmEF, 10.0 (5.0-20.2) for HFrEF, 8.8 (5.6-14.0) for unclassified HF]. Among participants without AF at baseline (n=5,761), 610 of them developed AF. Prevalent HF was associated with increased risk of AF [hazard ratio (95%CI) 2.3 (1.6-3.3) for HFpEF, 4.6 (2.4-8.6) for HFmEF, 3.8 (1.8-8.2) for HFrEF, 2.3 (0.9-5.6) for unclassified HR].

**Conclusion:** AF and HF frequently co-occur, with small differences by HF subtype, underscoring the importance of understanding the interplay of these two epidemics and evaluating shared preventive and therapeutic strategies.

## INTRODUCTION

Atrial fibrillation (AF) and heart failure (HF) are major public health problems, having emerged as growing epidemics in developed countries, the US included.^1^ The frequent co-existence of these two conditions portends significant morbidity, mortality and increased healthcare costs.^2^ It is estimated that by 2030, there will be over 12 million Americans with AF and 8 million with HF.^3, 4^ Almost two thirds of people with AF develop HF and one third of people with HF develop AF.^5^

Both HF and AF share common risk factors, pathophysiologic processes and adverse cardiovascular outcomes and, as such, are inextricably linked, with each disease predisposing to the other. Older age, smoking, obesity, hypertension, diabetes, coronary heart disease and renal disease are commonly shared predisposing factors to these conditions.^5-7^ Differences exist among HF subtypes in atrial remodeling and outcomes associated with AF.^8^ Similarly, clinical outcomes after AF are somewhat influenced by HF subtypes.^9^

Previous studies have explored the association between AF and HF. Most of these studies, however, are in hospital settings, which limit the understanding of the sequence of occurrence of both conditions in relation to the other.^10^ A study done in a large community cohort – the Framingham Heart Study – was comprised mostly of white participants, limiting its generalizability to other racial groups.^5^ Also, the Framingham analysis could not differentiate the association between different HF subtypes and incident AF due to limited sample size.

Given gaps in the existing literature, we sought to investigate the association of HF subtypes [HF with reduced ejection fraction (HFrEF), HF with mid-range ejection fraction (HFmEF) and HF with preserved ejection fraction (HFpEF)] with the incidence and prevalence of AF in a large community-based racially diverse cohort wherein the temporality of AF and HF can be more accurately obtained.

## METHODS

### Study population and setting

The ARIC study is a long term, prospective community-based study of the etiology of atherosclerosis and cardiovascular diseases. Detailed study methods have been previously described elsewhere.^11^ Briefly, the study started in 1987, and enrolled 15,792 participants aged 45 to 64 years (55.2% women) from 4 communities in the United States: Forsyth County, North Carolina; Jackson, Mississippi; Washington County, Maryland and northwest suburbs of Minneapolis, Minnesota. Participants were predominantly White in Washington County and Minneapolis, solely Black in Jackson and Black and White in Forsyth County. Following the baseline study visit (1987-1989), subsequent follow-up of participant has comprised of six clinical examinations (1990-1992, 1993-1995, 1996-1998, 2011-2013, 2016-2017, 2018-2019) and annual check-in phone calls to collect information on vital status and hospitalizations (semiannual starting in 2012). In addition, through regular surveillance of area hospitals, the ARIC study abstracts and reviews hospitalization data of its participants. The study was approved by the institutional review boards at participating centers and written consents were obtained from each participant.

For this analysis, we included only participants that attended the fifth study visit (2011-2013) because this was the earliest study time participants echocardiograms were taken in order to determine the ejection fraction. Of the 15,792 individuals recruited at baseline, 6,538 participated in the fifth visit. We excluded all participants who were of races other than White or African American from all sites as well as African Americans from the Minnesota and Washington county sites due to small sample size of these groups. In addition, participants whose AF status was not established at baseline were excluded.

### HF Ascertainment

Prevalent HF at study baseline (visit 5) was defined by the presence of at least one of the following at the time of the visit: a physician adjudicated diagnosis of HF, hospitalization with an International Classification of Diseases Ninth Revision, Clinical Modification (ICD-9-CM) discharge code of 428.x in the first position not overruled by a physician, self-reported HF or self-report of use of HF medication with pro-BNP greater than 125 pg/mL.^12^ These criteria have been previously validated.^13^ HF subtypes were determined based on left ventricular ejection fraction (LVEF) obtained by echocardiogram at study baseline and categorized using the 2016 European Society of Cardiology classification.^14^ Details about the echocardiographic protocol have been published elsewhere.^15^ Briefly, upon arrival at the center, participants were required to rest for 5 minutes, followed by blood pressure measurements after which the electrocardiography leads were positioned on the patient by trained sonographers. Depending on whether the subject was in sinus rhythm or had AF, at least 3 full cardiac cycles for each view or at least one 5 second acquisition per view were recorded respectively. The apical 4 and 2 chamber views were used to calculate left ventricular volumes by the modified Simpson’s method. LVEF was obtained from these volumes using standard calculations. All measurements were performed at the Echocardiography Reading Center (ERC; Brigham and Women’s Hospital, Boston, MA). HFpEF was defined by LVEF ≥ 50%, HFmEF was LVEF 40% - 49% and HFrEF was LVEF < 40%. Participants with prevalent HF as per prior definition but no information on LVEF were categorized as ‘unclassified’ HF.

### AF Ascertainment

The study outcome, AF, was determined using one of three methods: evidence of AF on a standard supine 12-lead resting ECG, hospitalization with an ICD-9-CM 427.31 or 427.32 or ICD-10-CM I48.x (starting in 2015) discharge code and if underlying cause of death was AF (ICD-10 code I48 or ICD-9 code 427.3). Standard 10-second 12-lead ECG recordings of study participants were done at baseline and at the subsequent study examinations using MAC PC Personal Cardiographs (Marquette Electronics Inc, Milwaukee, WI). Tracings were transmitted electronically to the ARIC ECG Reading Center (Epidemiological Cardiology Research Center, Wake Forest School of Medicine, Winston Salem, North Carolina) for automated reading, coding and storage. A cardiologist visually reviewed and confirmed AF diagnosis in tracings that had been automatically diagnosed as AF diagnosis. Using hospital discharge records, a trained abstractor identified and recorded International Classification of Diseases, Ninth Revision, Clinical Modification (ICD-9-CM) hospital discharge diagnosis AF was defined if ICD-9-CM codes 427.31 or 427.32 or ICD-10-CM I48.x were present in the absence of procedure codes for open heart surgery. A validity study of AF diagnosis using the aforementioned methods in the ARIC cohort showed a positive predictive value of ∼ 90%.^16^ The incidence date of AF was defined as the first occurrence of AF in an ECG during follow-up examination, a hospital discharge record, or death by AF whichever one occurred first. For the prevalent AF analysis, the outcome was defined as AF between visit 1 and visit 5. The incident AF analysis defined the outcome as new onset AF after visit 5 (2011 to 2013) through the end of 2017.

### Covariates

Covariates were obtained from visit 5 and included age, sex, race, study site, body mass index, systolic and diastolic blood pressure, smoking history, use of antihypertensive medication, diabetes mellitus, other cardiovascular diseases (coronary heart disease) and estimated glomerular filtration rate, which are known risk factors for AF.^6, 17^ Age, sex, race, and smoking status were self-reported. Height and weight were measured by technicians and the body mass index (BMI, in kg/m^2^) was calculated as weight (in kilograms) divided by height (in meters) squared. Blood pressure was measured three times using a standard protocol, and systolic and diastolic blood pressure was calculated as the average of the last two measurements. Diabetes was defined by one of the following criteria: 1) fasting glucose ≥ 126 mg/dL or non-fasting glucose ≥ 200 mg/dL, 2) self-reported physician diagnosis of diabetes, or 3) currently taking medication for diabetes. Coronary heart disease was defined as a history of myocardial infarction, coronary revascularization procedure or coronary artery bypass surgery, or the development of any of these during follow-up. Estimated glomerular filtration rate was calculated using the CKD-EPI equation for cystatin C.^18^

### Statistical analysis

Mean values with corresponding standard deviations and proportions were used to describe the baseline characteristics. The baseline prevalence of AF among patients with prevalent HF was calculated. To relate prevalent AF and prevalent HF subtypes, multivariable logistic regressions were used to examine the association between HF subtypes and prevalent AF first adjusting for demographic factors (age, sex, race) and then further adjusting for study site, BMI, systolic and diastolic blood pressure, smoking history, use of antihypertensive medication, diabetes mellitus, coronary heart disease and estimated glomerular filtration rate.

The sample used for incident AF analysis was free of AF at baseline. The cumulative incidence of AF was estimated using methods that consider death as a competing risk.^19^ A separate curve was used for those with and without HF, and by type of HF for incident AF. In order to assess the relationship between HF subtypes and incident AF, multivariable Cox proportional hazard models were fitted first adjusting for age, sex, and race and then further adjusting for study site, BMI, systolic and diastolic blood pressure, smoking history, use of antihypertensive medication, diabetes mellitus, coronary heart disease and estimated glomerular filtration rate. For both models, we assessed whether sex and race modified the association of AF with HF subtypes by fitting appropriate interaction terms to the models. Assumptions of collinearity and proportional hazard assumptions were tested.

Of the 6496 observations included in the analyses, there were none missing exposure nor outcome variables. Seventeen percent had at least one missing covariate. Missing data were assumed to be missing at random and imputed with the use of multivariate multiple imputation by chained equations (fully conditional specification, FCS) in order to reduce selection bias.^20^ We used the FCS method in order to specify different imputation models since we had both categorical and continuous variables. Variables included in the imputation model included exposure, outcome and covariates. Using PROC MI in SAS, 20 datasets were generated for multivariable model analyses and the results were consolidated to estimate regression parameters per Rubin’s formula using PROC MIANALYZE.^21^

In order to assess the impact of missing data, we did a complete case-only analysis as a sensitivity analysis, which restricted the analysis to participants with complete data on exposure, outcome, and confounder variables. Results with and without multiple imputation were not meaningfully different so we presented the former. Statistical analyses were performed in SAS version 9.4, and STATA version 16 (StataCorp LP, College Station, Texas). Two-tailed p-value of less than 0.05 was regarded as statistically significant.

## RESULTS

Of the 15,792 individuals recruited at baseline, 6,538 participated in the fifth study visit. After excluding participants who were of races other than White or African American from all sites as well as African Americans from the Minnesota and Washington county sites, there were 6,496 participants included in our final sample (Figure 1). Baseline characteristics of study participants, stratified by HF subtypes are shown in Table 1. The average age was 75.8 years (SD = 5.3). Most participants were female n (%) =3824 (58.9%), white 4977 (76.6%), and, on average, overweight (mean ± SD = 28.8±5.8). The mean systolic and diastolic blood pressures were 130.7 (SD = 18.7) and 66.3 (SD = 10.8) respectively. Participants with HF of any subtype were more likely to be smokers, consume alcohol, have diabetes, use antihypertensive medication, have coronary heart disease, have myocardial infarction and a lower estimated glomerular filtration rate (Table 1).

**Table 1.**
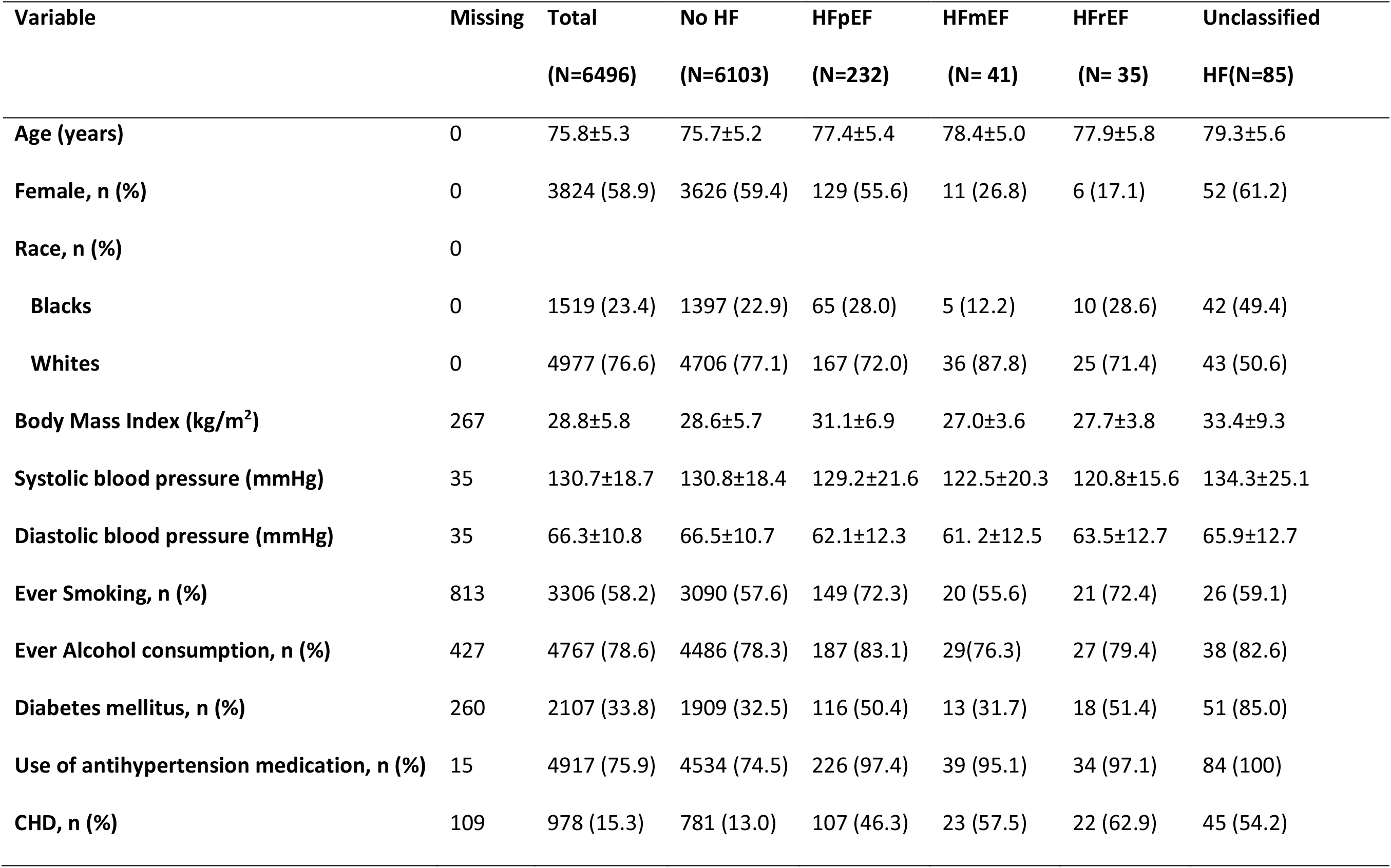
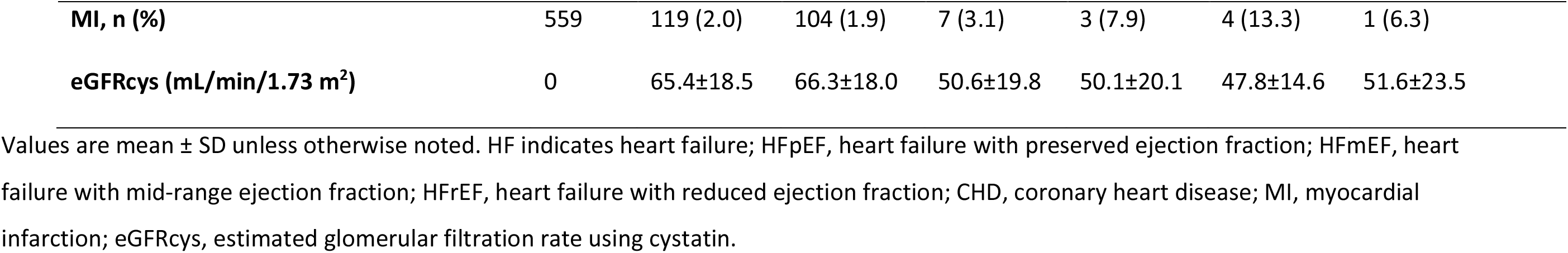
Sociodemographic and Clinical Characteristics of Participants in the ARIC Study by Heart Failure Subtype, 2011-2013.

**Figure 1.**
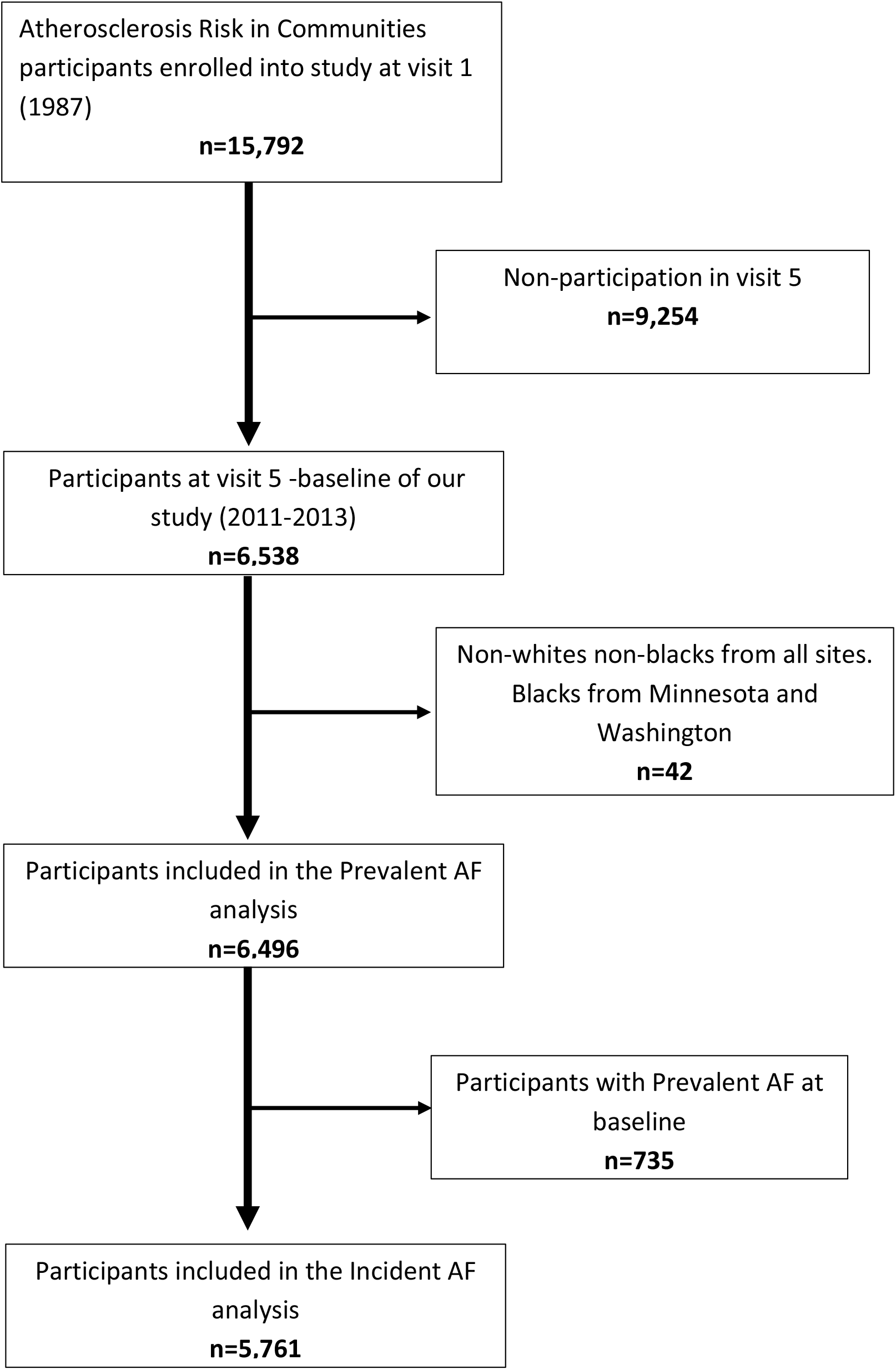
Study flow-chart of participants, ARIC 2011 to 2017

### Association of Prevalent HF subtypes with Prevalent AF

Among the 6,496 participants in the study, 735 of them had prevalent AF at baseline. The crude prevalence of AF was 43.5%, 51.2%, 54.3%, 48.2% and 9.1% among participants with HFpEF, HFmEF, HFrEF, unclassified HF and no HF categories respectively. Adjusting for age, sex and race as shown in logistic regression model 1 (table 2), compared to no HF, all HF subtypes were more likely to have prevalent AF (model 1, HFpEF, OR (95%CI): 7.4 (5.6-9.9); HFmEF OR (95%CI): 8.1 (4.3-15.3); HFrEF, OR (95%CI): 10.0 (5.0-20.2); unclassified HF, OR (95%CI): 8.8 (5.6-14.0)). Further adjustment yielded lower but more precise estimates. Compared to no HF, HF subtypes had 6 to 7 times the odds of prevalent AF controlling for age, sex, race, study site, BMI, systolic and diastolic blood pressure, smoking history, use of antihypertensive medication, diabetes mellitus, coronary heart disease and estimated glomerular filtration rate (model 2, HFpEF, OR (95%CI): 5.7 (4.2-7.7); HFmEF OR (95%CI): 6.3 (3.3-12.2); HFrEF, OR (95%CI): 6.8 (3.3-14.2); unclassified HF, OR (95%CI): 6.0 (3.7-9.7)). This association was not modified by sex (p > 0.05) and race (p >0.05).

**Table 2.** Association of Prevalent Atrial Fibrillation and Prevalent Heart Failure Subtypes, ARIC, 2011-2013

|  | No HF | HFpEF | HFmEF | HFrEF | Unclassified HF |
| --- | --- | --- | --- | --- | --- |
| **N** | 6103 | 232 | 41 | 35 | 85 |
| **AF prevalence \*** | 553 (9.1) | 101 (43.5) | 21 (51.2) | 19 (54.3) | 41 (48.2) |
| **Model 1†** | 1 (ref) | 7.4 (5.6-9.9) | 8.1 (4.3-15.3) | 10.0 (5.0-20.2) | 8.8 (5.6-14.0) |
| **Model 2 ‡** | 1 (ref) | 5.7 (4.2-7.7) | 6.3 (3.3-12.2) | 6.8 (3.3-14.2) | 6.0 (3.7-9.7) |

HF indicates Heart Failure; HFpEF, Heart Failure with preserved ejection fraction; HFmEF, Heart
Failure with mid-range ejection fraction; HFrEF, Heart Failure with reduced ejection fraction;
All values unless otherwise stated are odds ratios (95% confidence intervals)
\* Crude prevalence, n (%)
† Model 1: Logistic regression model adjusted for age, sex and race
‡ Model 2: Model 1 additionally adjusted for study site, BMI, systolic and diastolic blood pressure, smoking history, use of antihypertensive medication, diabetes mellitus, coronary heart disease and estimated glomerular filtration rate.

### Association of Prevalent HF subtypes with Incident AF

During a total of 29,023 person-years of follow-up (mean follow-up 4.4 years), we identified 610 incident cases of AF out of the 5,761 participants free of AF at baseline. The cases were distributed as follows by HF subtype: 36 with HFpEF, 9 with HFmEF, 6 with HFrEF, 9 with unclassified HF and 550 with no HF. The crude incidence rates of AF were 67.1, 121.6, 106, 66.5, 19.5 per 1000-person years among those with HFpEF, HFmEF, HFrEF, unclassified HF and no HF respectively (table 3). Cumulative incidence curves of AF by prevalent HF and HF subtypes accounting for competing risk of death are shown in Figures 2 and 3. The curve shows a higher risk of AF among those with HFrEF and HFmEF.

**Table 3.** Hazard ratios (95% confidence interval) of atrial fibrillation by heart failure subtypes, ARIC, 2011-2017

|  | No HF | HFpEF | HFmEF | HFrEF | Unclassified<br>HF |
| --- | --- | --- | --- | --- | --- |
| **AF cases** | 550 | 36 | 9 | 6 | 9 |
| **Person-years** | 28220.6 | 536.5 | 74 | 56.6 | 135.3 |
| **AF incidence \*** | 19.5 | 67.1 | 121.6 | 106.0 | 66.5 |
| **Model 1 †** | 1 (ref) | 3.4 (2.4-4.7) | 5.7 (3.1-10.3) | 5.0 (2.3-10.5) | 3.0 (1.5-5.7) |
| **Model 2 ‡** | 1 (ref) | 2.3 (1.6-3.3) | 4.6 (2.4-8.6) | 3.8 (1.8-8.2) | 2.3 (0.9-5.6) |

HF indicates Heart Failure; HFpEF, Heart Failure with preserved ejection fraction; HFmEF, Heart
Failure with mid-range ejection fraction; HFrEF, Heart Failure with reduced ejection fraction;
All values unless otherwise stated are hazard ratios (95% confidence intervals)
\* Crude incidence per 1000 person-years.
† Model 1: Cox proportional hazard model adjusted for age, sex and race

**Figure 2.**
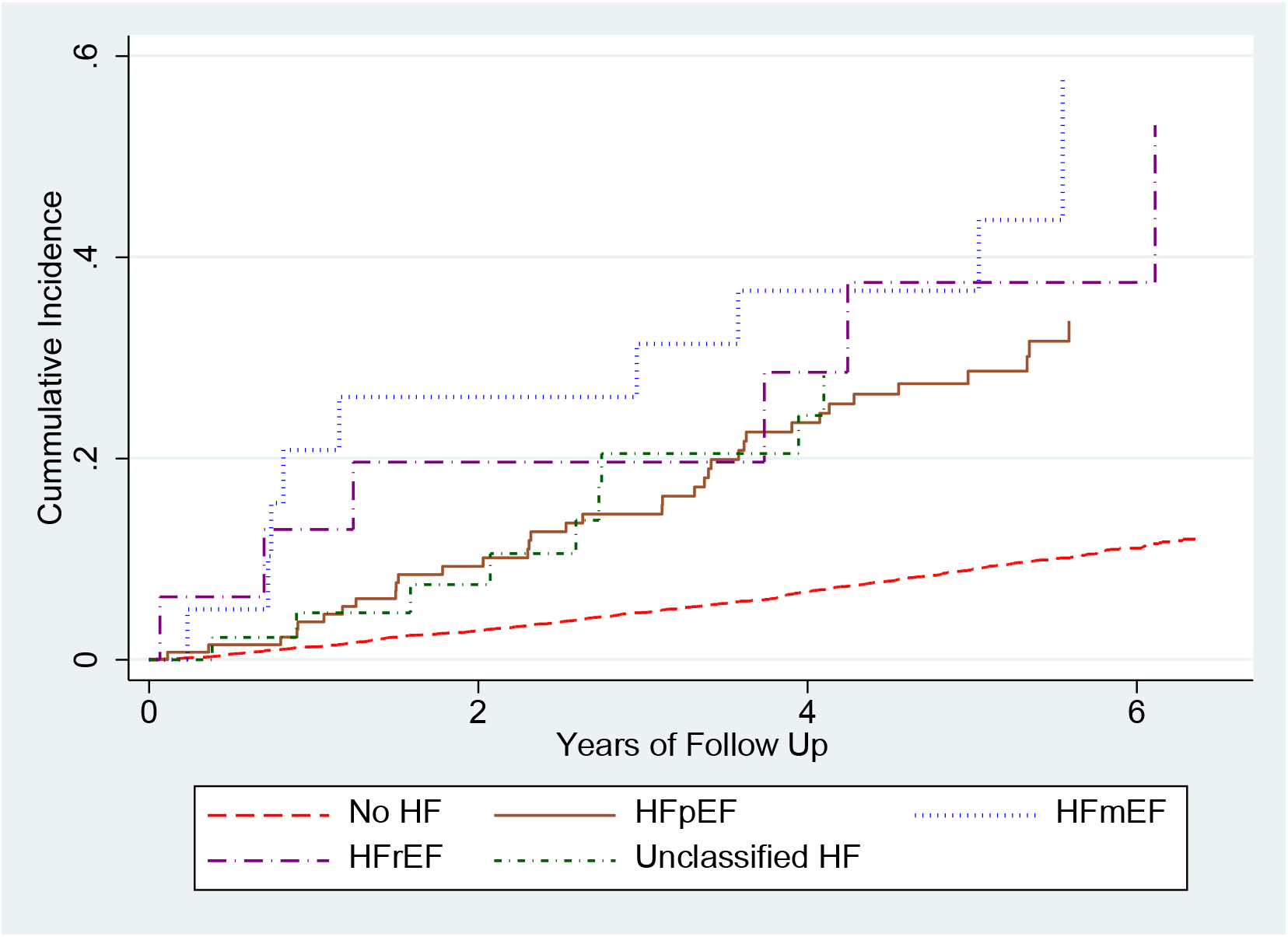
Cumulative incidence of AF considering death as competing risk among those without HF, HFpEF, HFrEF and Unclassified HF, ARIC cohort 2011 to 2017. HF, heart failure; HFpEF, heart failure with preserved ejection fraction; HFrEF, heart failure with reduced ejection fraction

**Figure 3.**
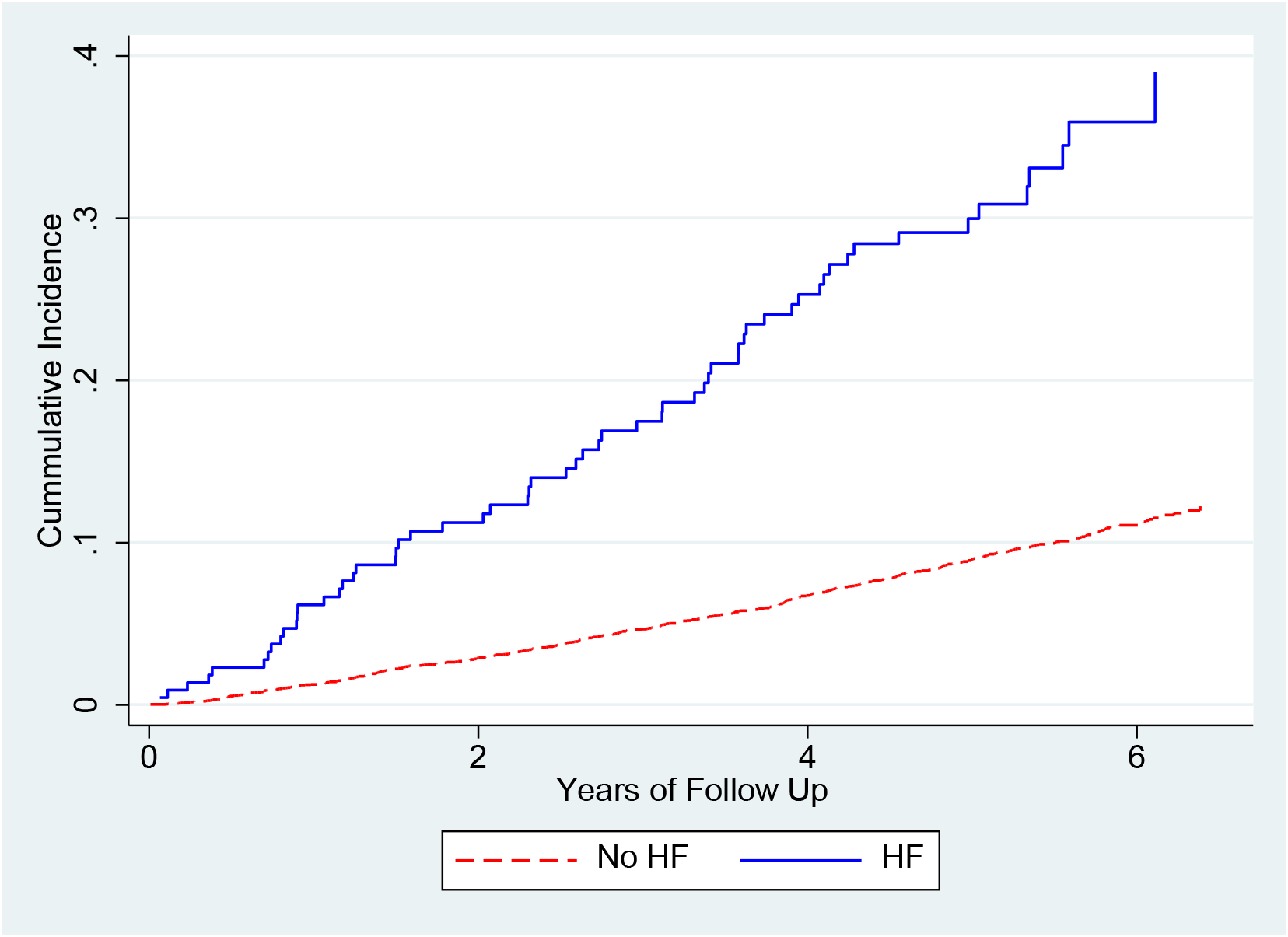
Cumulative incidence of AF considering death as competing risk among those with and without HF, ARIC cohort 2011 to 2017. HF, heart failure

The association between incident AF and prevalent HF subtypes is shown in table 3. Compared to no HF, HF subtypes were associated with a higher risk or incident AF adjusting for age, sex and race, (model 1, HFpEF, HR (95%CI): 3.4 (2.4-4.7); HFmEF, HR (95%CI): 5.7 (3.1-10.3); HFrEF, HR (95%CI): 5.0 (3.2-10.5); unclassified HF, HR (95%CI): 3.0 (1.5-5.7)). Further adjusting for study site, BMI, systolic and diastolic blood pressure, smoking history, use of antihypertensive medication, diabetes mellitus, coronary heart disease and estimated glomerular filtration rate reduced the association and improved precision, with all HF subtypes associated with 2-5 times higher risk of incident AF (model 2 HFpEF, HR (95%CI): 2.3 (1.6-3.3); HFmEF, HR (95%CI): 4.6(2.4-8.6) HFrEF, HR (95%CI): 3.8 (1.8-8.2); unclassified HF, HR (95%CI): 2.3 (0.9-5.6)). These associations were similar for both whites and blacks (p > 0.05) as well as men and women (p >0.05).

## DISCUSSION

Heart failure is a complex clinical syndrome with 3 subtypes based on LVEF-HFpEF, HFmEF, and HFrEF, all posing an increased independent risk of AF. Our analysis of the ARIC cohort showed that the prevalence and incidence of AF was high among HF patients of all subtypes, particularly among those with HFrEF. These findings highlight the co-occurrence of AF and HF. Heart failure, particularly HFrEF was associated with a 6 to 7 times increased odds of prevalent AF. Similarly, the risk of incident AF was 2-5 times higher in participants with HF.

According to previous studies, the prevalence of AF among those with HFrEF is high, ranging from 15% to 40%.^5, 22-25^ It appears to be even higher among participants with HFpEF reflecting their older age.^5, 22-24^ This is contrary to our findings where AF was slightly more common among participants with HFrEF, which could be due to the older age of ARIC participants compared to other studies. These discrepancies could also be as a result of the classification of HF subtypes being based on different cut points. Other studies defined HFrEF and HFpEF as LVEF ≤ 45% and > 45% respectively while our study used the modified ESC-HF criteria-LVEF of <40% to make a diagnosis of HFrEF and further distinguishing between those with midrange (LVEF = 40% - 49%) and preserved (LVEF ≥ 50%) ejection fractions. In a large community-based cohort study in Olmsted county, the incidence of AF among HFpEF was 31.6%, similar to our study incidence of 27.5%.^26^ However, in an analysis of a cohort of ambulatory patients with HF, the AF incidence was as low as 15%.^23^ This contrast could be due to differences between the studied populations, such as mean age, race and distribution of risk factors of AF.

The high prevalence and incidence of AF among participants with HF of any subtype, also previously demonstrated in large community-based Framingham and Olmsted cohorts, partly reflects the shared predisposing factors such as age, race, diabetes, hypertension and other cardiovascular diseases in both conditions. Furthermore, the pathophysiological processes underlying HF and AF are closely related. As a result of sustained increase in atrial pressure in persons with left ventricular dysfunction and overt HF, a process of atrial remodeling ensues which leads to atrial wall fibrosis, heterogeneity in conduction and impaired contraction of this atria. This eventually can lead to AF onset among patients with HF.

AF was present in 51.2% of participants with HFmEF at baseline. This prevalence is higher than previously reported estimates of 22% in ESC-HF registries,^25^ 20% in clinic data,^27^ or 40% in the American Heart Association national registries for HF.^22^ The higher prevalence in our study may be due to the older age of the participants and the rigorous ascertainment of AF cases, as opposed to passive ascertainment in registry data. There have been conflicting results on the similarity of HFmEF to HFpEF and HFrEF in terms of underlying comorbidities such AF. HFmEF has commonly been classified as HFpEF suggesting that both types share similarities in disease characteristics, management options and clinical outcomes.^22, 28^ Other studies have stipulated that HFmEF is rather similar to HFrEF and the underlying prevalence of AF in this group is not significantly different from that of HFrEF.^27^ In our study, those with HFmEF had the highest risk (122.6 per 1000 person-years) of developing AF. This unexpectedly high burden of AF in HFmEF emphasizes the need to study the underlying disease characteristics as a separate subgroup.^14^

This analysis has several strengths. To the best of our knowledge, our study is the first to study the association of AF in HF in an epidemiologically representative population-based cohort using the modified 2016 ESC classification which makes a further distinction in HF categories by defining an HFmEF subtype. In addition, while previous studies have been conducted in hospital settings, using smaller samples, and in predominantly white cohorts,^5, 23, 26, 29^ the ARIC cohort is a large community-based, racially and geographically diverse population consisting of whites and blacks from four communities in the United States. This allows for greater generalizability.

The results of our analysis should be interpreted in the context of some limitations. AF was ascertained using study electrocardiograms and hospital discharge records. This could lead to under ascertainment of AF cases diagnosed on outpatient basis as well as asymptomatic and paroxysmal AF cases. Nonetheless, the validity of using the aforementioned methods for AF ascertainment is satisfactory.^16, 30^ Twenty-two percent of HF cases were unclassified due to the absence of LVEF measurements which may have led to misclassification bias potentially influencing our results. Despite our large sample, the number of AF events in each HF category was limited, reducing the precision of our estimates of association. Because ARIC study visits comprise of comprehensive in-person exams, participants who complete the fifth visit are more likely to be healthier compared to those who do not attend this visit. This healthy-participant effect disproportionately affects participants with HFrEF who are sicker and more likely to miss a study visit. As a result of the observational nature of the study, we cannot rule out residual unmeasured confounding. Our participants were whites and blacks, limiting generalizability of our results to other racial groups.

In conclusion, in this community-based cohort, we show that participants with HF are more likely to have underlying AF or are at risk for incident AF. The frequent co-occurrence of the 2 conditions underscores the importance of the growing public health problem. Also, due to an unexpectedly high burden of HFmEF, it is vital for further studies to probe into this separate HF category to understand its disease characteristics.

## Data Availability

The data underlying this article cannot be shared publicly to protect the privacy of study participants. The data can be obtained from the ARIC Coordinating Center following procedures available in the following website: https://sites.cscc.unc.edu/aric/distribution-agreements

## ACKNOWLEDGMENT

The authors thank the staff and participants of the ARIC study for their important contributions.

